# Sequence Analysis of 20,453 SARS-CoV-2 Genomes from the Houston Metropolitan Area Identifies the Emergence and Widespread Distribution of Multiple Isolates of All Major Variants of Concern

**DOI:** 10.1101/2021.02.26.21252227

**Authors:** S. Wesley Long, Randall J. Olsen, Paul A. Christensen, Sishir Subedi, Robert Olson, James J. Davis, Matthew Ojeda Saavedra, Prasanti Yerramilli, Layne Pruitt, Kristina Reppond, Madison N. Shyer, Jessica Cambric, Ilya J. Finkelstein, Jimmy Gollihar, James M. Musser

## Abstract

Since the beginning of the SARS-CoV-2 pandemic, there has been international concern about the emergence of virus variants with mutations that increase transmissibility, enhance escape from the human immune response, or otherwise alter biologically important phenotypes. In late 2020, several “variants of concern” emerged globally, including the UK variant (B.1.1.7), South Africa variant (B.1.351), Brazil variants (P.1 and P.2), and two related California “variants of interest” (B.1.429 and B.1.427). These variants are believed to have enhanced transmissibility capacity. For the South Africa and Brazil variants, there is evidence that mutations in spike protein permit it to escape from some vaccines and therapeutic monoclonal antibodies. Based on our extensive genome sequencing program involving 20,453 virus specimens from COVID-19 patients dating from March 2020, we report identification of all important SARS-CoV-2 variants among Houston Methodist Hospital patients residing in the greater metropolitan area. Although these variants are currently at relatively low frequency in the population, they are geographically widespread. Houston is the first city in the United States to have all variants documented by genome sequencing. As vaccine deployment accelerates worldwide, increased genomic surveillance of SARS-CoV-2 is essential to understanding the presence and frequency of consequential variants and their patterns and trajectory of dissemination. This information is critical for medical and public health efforts to effectively address and mitigate this global crisis.

## [Introduction]

The severe acute respiratory syndrome coronavirus 2 (SARS-CoV-2) is the causative agent of coronavirus disease 2019 (COVID-19). Since first being identified in December 2019,^1-4^ the virus has spread globally and is responsible for massive human morbidity and mortality worldwide.^5-9^ At the onset of the pandemic, effective treatments for COVID-19 were lacking. But as a result of intense global research efforts, monoclonal antibody (mAbs) therapies^10, 11^ and several vaccines,^12, 13^ primarily directed against the spike protein, have been developed to treat and prevent SARS-CoV-2 infection.

In late 2020 the international research community described several SARS-CoV-2 “variants of concern” that warranted special scrutiny. These include the United Kingdom (UK) variant (B.1.1.7), South Africa variant (B.1.351), Brazil variants (P.1 and P.2) and two California variants (B.1.429/CAL.20C and B.1.427/CAL.20C).^14-22^ These virus variants were designated as “concerning” predominantly due to their reported enhanced person-to-person transmission in some geographic areas, and they have since been detected in several countries worldwide. For example, the UK B.1.1.7 variant spread rapidly in southeast England where it caused large numbers of COVID-19 cases,^14^ and was identified shortly thereafter in the United States (US) [CDC; https://www.cdc.gov/coronavirus/2019-ncov/transmission/variant.html]. ^23^ More than 1,600 cases have since been documented in the US, and at least one large outbreak recently was reported in a Michigan prison.^24, 25^ There is concern at the Centers for Disease Control and Prevention (CDC) that it could become the dominant variant causing disease in the US by March.^23, 24, 26^ Moreover, the UK B.1.1.7 variant may be linked to an increased death rate compared to other virus types, adding further concern.^18, 21, 27, 28^

Similarly, the South Africa and Brazil variants caused large disease outbreaks in their respective countries.^19, 20^ These variants also are of concern because they contain a mutation (E484K) in the spike protein that decreases efficacy of some therapeutic mAbs, decreases in vitro virus neutralization, and may result in potential escape from immunity induced by natural infection or vaccination.^29-37^ All three variants (UK B.1.1.7, Brazil P.1, and South Africa B.1.351) also have a N501Y mutation in spike protein that is associated with stronger binding to the ACE2 receptor, possibly contributing to increased transmissibility.^38,39^

The Houston metropolitan area is the fourth largest and most ethnically diverse city in the US, with a population of approximately 7 million (https://www.houston.org/houston-data).^40^ The 2,400-bed Houston Methodist health system has eight hospitals and cares for a large, multiethnic, and geographically and socioeconomically diverse patient population throughout greater Houston. The eight Houston Methodist hospitals have a single central molecular diagnostic laboratory, which means that all RT-PCR-specimens can readily be identified, banked, and subjected to further study as needed. In addition, the Department of Pathology and Genomic Medicine has a long-standing record of integrating genome sequencing efforts into clinical care and research, especially related to microbial pathogens infecting our patients.^41-49^ In the aggregate, strategic co-localization of these diagnostic attributes coupled with a contiguous research institute building seamlessly facilitates comprehensive population genomic studies of SARS-CoV-2 viruses causing infections in the Houston metropolitan region.^46,49^

Before the SARS-CoV-2 virus arrived in Houston, we planned an integrated strategy to confront and mitigate this microbial threat to our patients. In addition to rapidly validating an RT-PCR test for the virus, we instituted a plan to sequence the genome of every positive specimen from patients within the Houston Methodist system, with the goal of understanding pathogen spread in our community and identifying biologically-important mutant viruses. We previously described the detailed population genomics of the first and second waves of SARS-CoV-2 in the Houston metropolitan region.^46,49^ We have continued to sequence positive SARS-CoV-2 specimens with the goal of monitoring for variants of concern and genome mutations that may be associated with patient outcome or therapeutic failure.

This report describes the identification of multiple isolates of important SARS-CoV-2 variants, including the UK B.1.1.7, South Africa B.1.351, Brazil P.1 and P.2, and California B.1.429 and B.1.427 variants in Houston patient specimens collected from December 2020 through mid-February 2021. These findings represent the first detection of the South Africa and Brazil variants in Texas and only the second time UK variants have been identified in Houston. Greater Houston is the first metroplex in the US documented to have all of these important and concerning variants circulating among its residents. Our discoveries further illustrate the need for increased population genomic and epidemiology efforts to identify and help track dissemination of these variants, monitor development of new variants, and assess the relationship between variants and COVID-19 disease outcomes.

## Materials and Methods

### Patient Specimens

All specimens were obtained from individuals who were registered patients at Houston Methodist hospitals, associated facilities (e.g. urgent care centers), or institutions in the greater Houston metropolitan region that use our laboratory services. Virtually all individuals had signs or symptoms consistent with COVID-19 disease. This work was approved by the Houston Methodist Research Institute Institutional Review Board (IRB1010-0199).

### SARS-CoV-2 Molecular Diagnostic Testing

Specimens obtained from symptomatic patients with a high degree of suspicion for COVID-19 disease were tested in the Molecular Diagnostics Laboratory at Houston Methodist Hospital using assays granted Emergency Use Authorization (EUA) from the FDA (https://www.fda.gov/medical-devices/emergency-situations-medical-devices/faqs-diagnostic-testing-sars-cov-2#offeringtests). Multiple molecular testing platforms were used, including the COVID-19 test or RP2.1 test with BioFire Film Array instruments, the Xpert Xpress SARS-CoV-2 test using Cepheid GeneXpert Infinity or Cepheid GeneXpert Xpress IV instruments, the SARS-CoV-2 Assay using the Hologic Panther instrument, the Aptima SARS-CoV-2 Assay using the Hologic Panther Fusion system and the SARS-CoV-2 assay using Abbott Alinity m instruments. All assays were performed according to the manufacturer’s instructions. Testing was performed on material obtained from nasopharyngeal, oropharyngeal, or nasal swabs immersed in universal transport media (UTM), bronchoalveolar lavage fluid, or sputum treated with dithiothreitol (DTT). To standardize specimen collection, an instructional video was created for Houston Methodist healthcare workers (https://vimeo.com/396996468/2228335d56).

### SARS-CoV-2 Genome Sequencing

Libraries for whole virus genome sequencing were prepared according to version 3 of the ARTIC nCoV-2019 sequencing protocol (https://artic.network/ncov-2019). Long reads were generated with the LSK-109 sequencing kit, 24 native barcodes (NBD104 and NBD114 kits), and a GridION instrument (Oxford Nanopore). Short sequence reads were generated with either a NextSeq 550 or NovaSeq 6000 instrument (Illumina).

### SARS-CoV-2 Genome Sequence Analysis

Viral genomes were assembled with the BV-BRC SARS-Cov2 assembly service (https://www.bv-brc.org/app/ComprehensiveSARS2Analysis).^50^ The One Codex SARS-CoV-2 variant calling and consensus assembly pipeline was chosen for assembling all sequences (https://github.com/onecodex/sars-cov-2.git) using default parameters and a minimum read depth of 3. Briefly, the pipeline uses seqtk version 1.3-r116 for sequence trimming (https://github.com/lh3/seqtk.git); minimap version 2.1^51^ for aligning reads against reference genome Wuhan-Hu-1 (NC_045512.2); samtools version 1.11 for sequence and file manipulation^52^; and iVar version 1.2.2 for primer trimming and variant calling.^53^

### Geospatial Analysis

The patient home address zip codes were used to visualize the geospatial distribution of spread for each variant of concern. Figures were generated using Tableau version 2020.3.4.

## Results

Since the start of the SARS-CoV-2 pandemic, we have sequenced 20,453 specimens collected from patients in the Houston metropolitan area. In genome sequencing conducted in January and February 2021, we discovered our first variants of concern. These included 23 UK variants (B.1.1.7), two South African variants (B.1.351), and four Brazilian variants (P.1). We also identified 162 patients infected with the California variants (B.1.429, *N* = 143; B.1.427, *N* = 19) and 39 patients infected with Brazil P.2 variants 2020 (Table 1).

**Table 1.** Variants of concern or variant of interest identified in the Houston Metropolitan area.

| Variant | No. of Isolates |
| --- | --- |
| B.1.1.7 | 23 |
| B.1.351 | 2 |
| P.1 | 4 |
| P.2 | 39 |
| B.1.429 | 143 |
| B.1.427 | 19 |

### UK Variant of Concern (B.1.1.7)

The UK variant known as B.1.1.7 was first identified in September 2020 in the UK and was designated as a variant of concern in South London on December 14, 2020. It was strongly associated with a resurgence of SARS-CoV-2 infections in that region and rapidly became the dominant lineage.^26^ Importantly, the UK has the most extensive SARS-CoV-2 genome sequencing program in the world, making them particularly well situated to rapidly identify new variants. Of the ∼500,000 SARS-CoV-2 genome sequences submitted to GISAID from global sources, approximately one-half originated from collaborating laboratories in the UK as part of the COVID-19 Genomics UK Consortium.^54, 55^

The UK B.1.1.7 variant is of particular concern because it has an unusually large number of genome mutations, including multiple changes in spike protein (Figure 1). Some of the mutations of primary concern include N501Y located in the receptor binding domain, and a two amino acid deletion (del69-70) that has arisen in multiple SARS-CoV-2 genetic backgrounds and is associated with increased transmissibility^26^. In addition, evidence has been presented from the UK that B.1.1.7 strains may cause increased hospitalization and mortality.^18, 21, 27, 56^ The first patient we identified in Houston with a B.1.1.7 variant was diagnosed the second week of January, 2020; thus far we have identified 23 patients with this variant of concern (Table 1). Of note, none of our first three patients had an international travel history, suggesting that they acquired the B.1.1.7 infections either locally or during domestic travel. Preliminary evidence indicates that immune sera from the Pfizer-BioNTech SARS-CoV-2 vaccine retain the ability to neutralize B.1.1.7 variants *in vitro*.^57^ Additional studies have found that convalescent plasma from many patients, and some monoclonal antibody therapies, retain the ability to neutralize B.1.1.7 variant SARS-CoV-2 *in vitro*.^34, 35^

**Figure 1.**
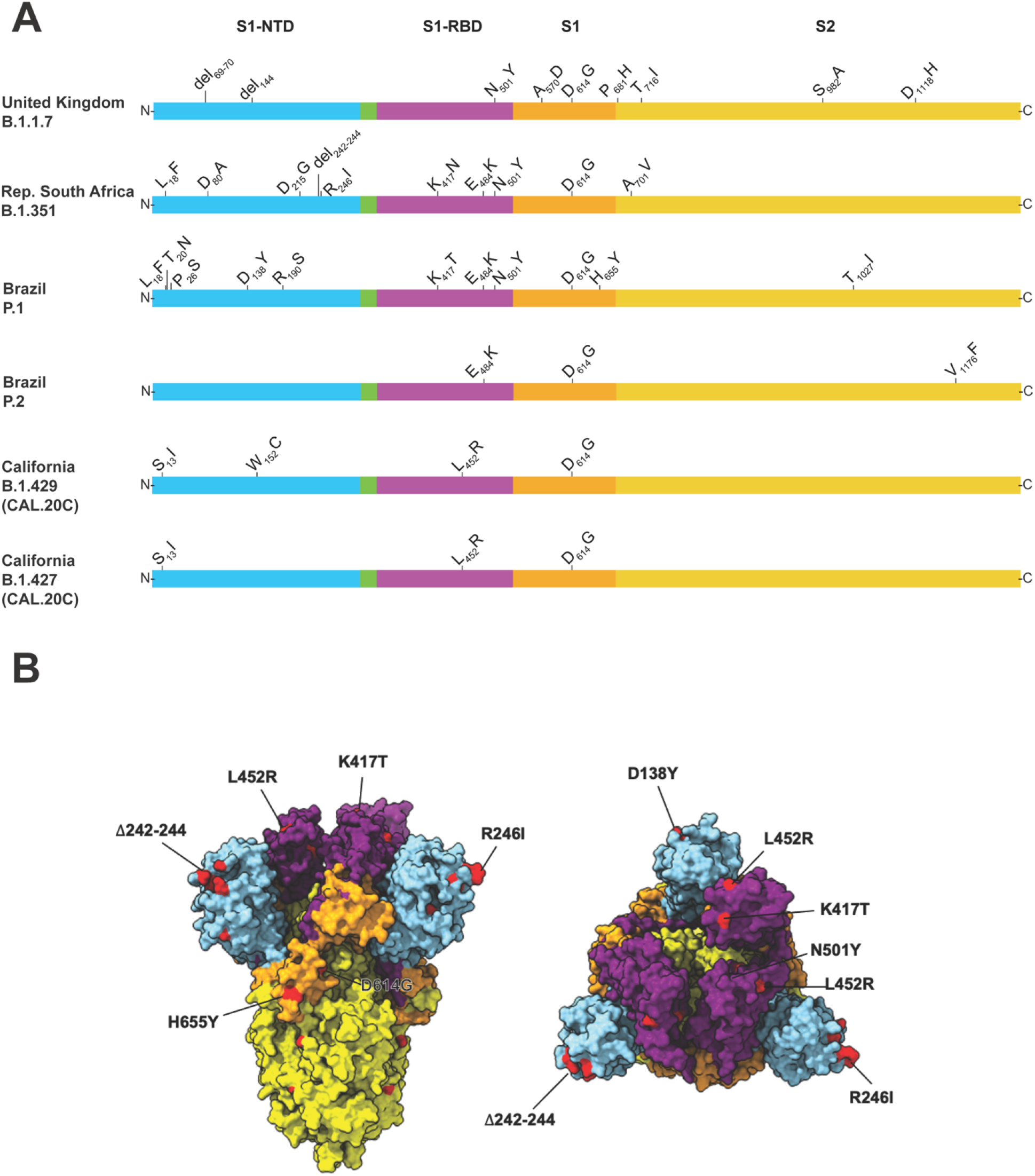
**A**: Schematic showing structural changes present in the spike protein of the major SARS.CoV.2 variants identified in the study. S1-NTD, S1 domain-aminoterminal domain; S1-RBD, S1 domain-receptor binding domain; S1, S1 domain; S2, S2 domain. B: Mapping of important changes onto the cryoEM structure of spike protein. The color scheme matches that used in panel A. Blue (NTD), purple (RBD), orange (S1), and yellow (S2). Aggregate mutations present in variants of concern are colored in red when amino acid residues are present in the resolved structure. Left, side view of SARS-CoV-2 prefusion-stabilized spike. Right, top view. Structure of PDB 6vsb was used as reference.

### South Africa Variant of Concern (B.1.351)

The South Africa B.1.351 variant of concern was first identified in a COVID-19 epidemic wave occurring in Nelson Mandela Bay in October 2020.^19^ This variant was concerning because of its large number of spike protein mutations (including K417N, E484K, and N501Y) (Figure 1) and apparent increased transmissibility.^19, 38^ These three mutations are located in the receptor binding domain of spike and may decrease the effectiveness of some mAb therapies and vaccines.^29-31, 34, 35, 58^ The first South Africa variant detected in Houston was identified in a patient specimen we collected the last week of December, 2020, and the second patient’s specimen was collected the first week of January, 2021. Of note, these Houston Methodist Hospital patients had no known international travel history, suggesting domestic acquisition of this B.1.351 variant.

### Brazil Variants of Concern (P.1 and P.2)

The P.1 variant of concern was reported to have originated in Manaus, Brazil, and like the South Africa B.1.351 variant, has numerous mutations in spike protein, including E484K and N501Y (Figure 1).^59^ We identified our first P.1 variant in Houston specimens the third week of January, 2021. In total, we have identified four P.1 variants in our patient samples (Table 1). The P.2 variant began to spread in Brazil in earnest in October of 2020, similar to P.1.^60, 61^ It also has a E484K amino acid change in the RBD of spike protein (Figure 1), similar to variant P.1 and B.1.351.^17^ We first identified a P.2 variant in a patient specimen obtained the last week of December, 2020. In total, we have documented 39 P.2 variants in our patient specimens (Table 1).

### California Variants (B.1.429 and B.1.427)

The emergence of what became known as the California variant, originally known as CAL.20C and later designated as lineages B.1.429 and B.1.427, was first identified in Los Angeles County in July 2020 as a single isolate.^62, 63^ This variant re-emerged in October 2020 and was associated with an increasing number of cases during a wave of SARS-CoV-2 infections in the region.^16^ Variant B.1.429 accounted for 36% of isolates collected from late November to late December 2020 in Los Angeles County.^16^ Since November 2020, this variant has been detected in 42 states in the US,^63^ and was first found in Houston Methodist Hospital patients in specimens obtained the last week of December, 2020. We identified 143 and 19 patients with the B.1.429 and B.1.427 isolates, respectively (Table 1). The B.1.427 variant is closely related to B.1.429 (Figure 1) and has spread from California to 34 states since October 2020.^62^ The California variants are noteworthy primarily for their emergence and very rapid spread in Los Angeles County and identification elsewhere in the US. However, as of February 17, 2021, they have not been designated as variants of concern by the Centers for Disease Control.

### Geospatial Distribution of Variants

Given the importance of the identification of these SARS-CoV-2 variants in the Houston metropolitan area, we examined their geospatial distribution to investigate the extent of dissemination (Figure 2). With the exception of the B.1.351 variant, patients infected with all other variants resided in widely dispersed areas of the metropolitan area. This finding is consistent with the well-known propensity of SARS-CoV-2 to spread rapidly between individuals, and especially so for these variants of concern ^19, 23, 24, 27, 64-66^.

**Figure 2.**
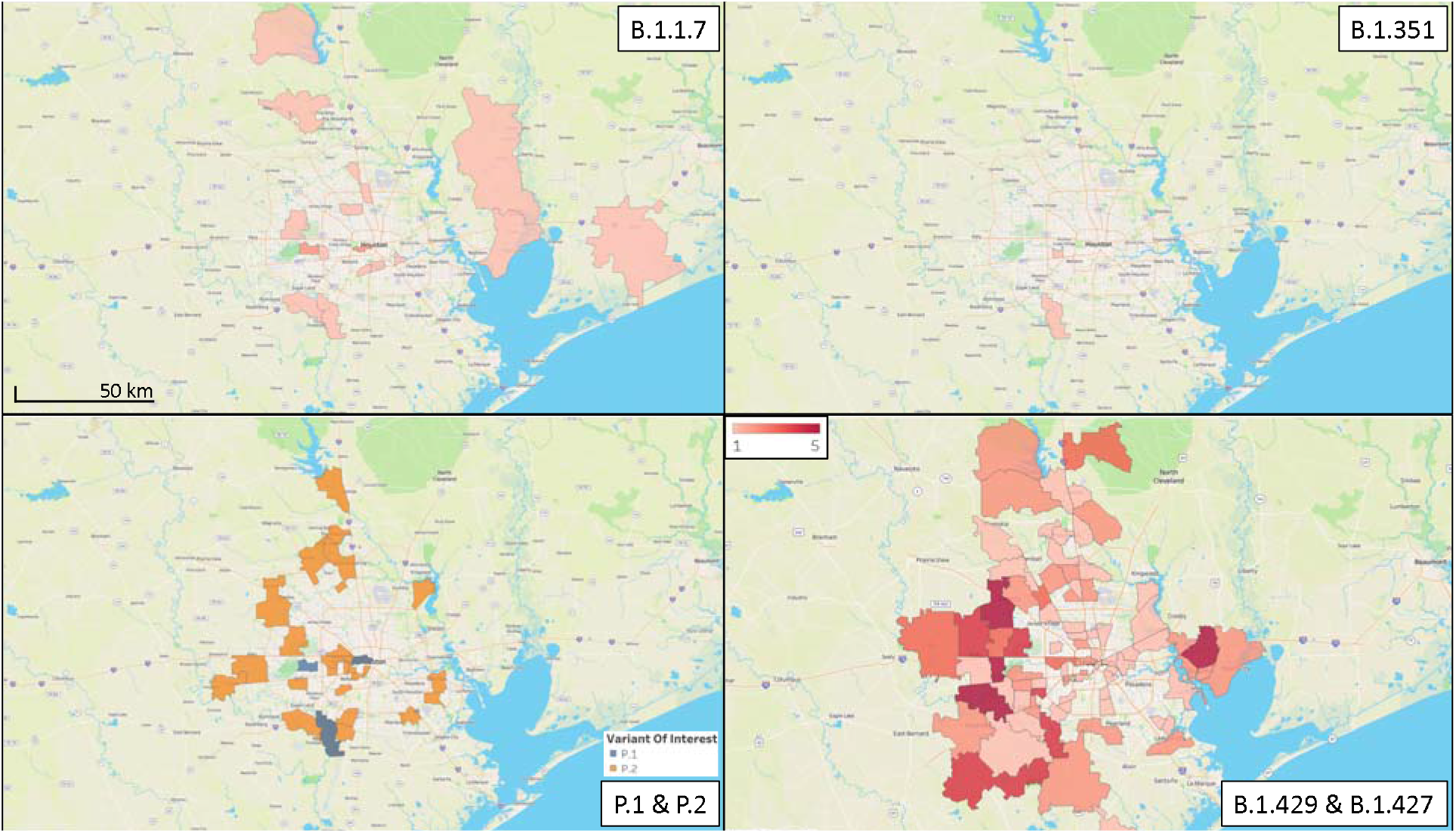
Geospatial distribution for each variant of concern identified in the study. The home address zip code for each patient was used and figures were generated using Tableau version 2020.3.4.

## Discussion

Here we report discovery of the UK (B.1.1.7), South Africa (B.1.351), and Brazil (P.1) SARS-CoV-2 variants of concern from patients in the Houston metropolitan region. We also identified geographically-widespread dissemination of the Cal.20C California (B.1.429 and B.1.427) variants of interest. These four SARS-CoV-2 variants are distributed across a large geospatial region in the metropolitan region (Figure 2), indicating successful patient-to-patient transmission among Houstonians. None of the affected patients were from a common household or reported recent international travel, suggesting that every infection was independently acquired locally or during domestic travel. Given that Houston is a culturally- and ethnically-diverse population center with two international airports, a major shipping center, and a global energy sector, the discovery of patients infected with each of the four concerning SARS-CoV-2 variants is not unexpected but it is disquieting. With this report, Houston now becomes the first US city to document patients infected with each of the four SARS-CoV-2 variants of concern or interest, testament to our aggressive sequencing of COVID-19 patient samples.

The P.2 variant gained recent attention in the scientific and lay press because it has been reported to cause SARS-CoV-2 reinfections.^67, 68^ We identified 39 P.2 infections among Houston patients. Although it is currently a numerically minor cause of all Houston-area infections, P.2 is now the most common SARS-CoV-2 variant of concern in our population.

The E484K amino acid replacement in spike protein is characteristic of P.1, P.2, and B.1.351 strains (Figure 1). It has independently arisen in many different SARS-CoV-2 genomic backgrounds, including some B.1.1.7 strains.^69^ This amino acid replacement has caused substantial public health concern due to its potentially detrimental effects on neutralizing activity of therapeutic mAbs, sera obtained from naturally infected individuals, and post-vaccination sera.^70, 71^ That is, the E484K amino acid change may facilitate vaccine escape. Among our Houston SARS-CoV-2 genomes, E484K was detected 84 times (0.4% of the total genomes sequenced). It was first detected in a respiratory specimen collected in July 2020, near the peak of our second massive wave of infections,^46^ and has been identified in many diverse genomic backgrounds thereafter. Due to this strong signal of convergent evolution, we will continue to closely monitor all Houston SARS-CoV-2 genomes for the E484K amino acid change.

Recently, the Q677H amino acid change in spike protein has been identified in SARS-CoV-2 patient samples collected in multiple US states and other global locations.^72, 73^ Q677H has arisen in at least six distinct genomic backgrounds.^73^ A Q667P amino acid change has also been identified.^73^ Among the Houston genomes, Q677H occurred 288 times (1.4%) and is encoded by two different nucleotide changes. We also identifed two other amino acid changes, 677P (in 330 genomes, 1.6%) and Q677K (2 genomes, <0.1%) in Houston. Taken together, these data suggest selection for a yet to be determined biologic phenotype associated with amino acid replacements at position 677.

Many population genomic studies performed in varous global locations have clearly demonstrated that SARS-CoV-2 variants with biologically-relevant phenotypes have evolved. Emergence of new variants underscores the need for ongoing extensive genomic sequencing efforts for early identification and public health warning. In support of these efforts, our laboratory has devoted substantial resources to SARS-CoV-2 genomics, resulting in sequence analysis of more genomes than any other state in the US.^54^ Since March 2020, approximately 36,500 SARS-CoV-2 positive patients have received care in our Houston Methodist health system, and we have sequenced 20,453 virus genomes. In total, this dataset represents 56% of our Houston Methodist COVID-19 patients. Inasmuch as almost 500,000 COVID-19 infections have been reported in the Houston metropolitan area,^74^ we have sequenced the genome of 4.1% of all cases reported in our area. Based on modeling, this sample depth may be sufficient to identify all variants occurring at a biologically-relevant frequency.^75^ Due to the very wide geographic catchment of our eight-hospital system that serves a very diverse patient population, the data presented here likely reflect a reasonably detailed overview of SARS-CoV-2 genomic diversity throughout our metroplex. This comparatively deep sampling of the Houston metropolitan SARS-CoV-2 population enabled us to identify patients infected with variants of concern, and provided information regarding the timeframe of initial presence and frequency of each variant. We modeled our strategy on the aggressive genome sequencing being conducted in the UK, a global leader in SARS-CoV-2 genome sequencing.^76^

Our large SARS-CoV-2 genome dataset and comprehensive infrastructure are unique resources. By linking the SARS-CoV-2 whole genome sequence data to patient metadata present in our electronic medical record, we are able to use analytic tools such as high-performance compute clusters and machine learning to investigate the relationship between genomic diversity and phenotypic traits such as strain virulence or patient outcomes.^46^ For example, recent reports of increased mortality caused by B.1.1.7 variant strains are very concerning and worthy of further investigation.^18, 21, 27, 28^ Similarly, our COVID-19 biobank has cryopreserved respiratory samples, white blood cells, serum, plasma, and formalin-fixed paraffin-embedded tissues for use in downstream investigations such as viral neutralization assays, RNA sequencing, and immune repertoire analysis.

Our goal is to sequence the SARS-CoV-2 genome of every infected patient in our health care system in near-real time, and expand outward to other patients in our community. Consistent with these goals, the American Rescue Plan announced by the Biden administration proposes to substantially fund sequencing capacity in the US. However, it remains unclear how these important funds will be distributed.^77^ Our results from a major metropolitan region in the US underscore the necessity of greatly increased genome surveillance to rapidly identify and track the emergence and introduction of SARS-CoV-2 variants in the US and local areas.

## Data Availability

All genomes have been submitted to GISAID.

https://www.gisaid.org/

## Acknowledgments

We thank Drs. Jessica Thomas and Zejuan Li, Erika Walker, the many very talented and dedicated molecular technologists, and many labor pool volunteers in the Molecular Diagnostics Laboratory and Methodist Research Institute for their dedicated efforts. We are indebted to Drs. Marc Boom and Dirk Sostman for their support, to many very generous Houston philanthropists for their tremendous support and to the Houston Methodist Academic Institute Infectious Diseases Fund that have made this ongoing project possible. James Davis and Robert Olson were funded in whole or in part with Federal funds from the National Institute of Allergy and Infectious Diseases, National Institutes of Health, Department of Health and Human Services, under Contract No. 75N93019C00076. We thank Jessica W. Podnar and personnel in the University of Texas Genome Sequencing and Analysis Facility for sequencing some of the genomes in this study. We acknowledge the assistance of Maulik Shukla and Marcus Nguyen at PATRIC Bioinformatics Resource Center for their assistance with sequence analysis. We gratefully acknowledge the originating and submitting laboratories of the SARS-CoV-2 genome sequences from GISAID’s EpiFlu^™^ Database used in some of the work presented here. We also thank many colleagues for critical reading of the manuscript and suggesting improvements, and Dr. Kathryn Stockbauer, Sasha Pejerrey, Adrienne Winston, and Heather McConnell for help with figures, tables, and editorial contributions.

## Author Contributions

J.M.M. conceptualized and designed the project; S.W.L, R.J.O., P.A.C., S.S., R.O., J.J.D., M.S., P.Y., L.P., K.R., M.N.S, J.C., I.J.F, and J.G. performed research. All authors contributed to writing the manuscript.

## Data availability

All genomes have been submitted to GISAID (www.gisaid.org)

